# Breastfeeding mother-child clinical outcomes after COVID-19 vaccination

**DOI:** 10.1101/2021.06.19.21258892

**Authors:** Jia Ming Low, Le Ye Lee, Yvonne Peng Mei Ng, Youjia Zhong, Zubair Amin

**Affiliations:** Department of Paediatrics, Yong Loo Lin School of Medicine, National University of Singapore, Singapore; Department of Neonatology, Khoo Teck Puat-National University Children’s Medical Institute, National University Health System, Singapore; Khoo Teck Puat-National University Children’s Medical Institute, National University Health System, Singapore; Duke-NUS Medical School, Singapore

**Keywords:** COVID-19, BNT162b2, mRNA vaccines, breastfeeding, human milk, mastitis

## Abstract

This is a prospective cohort study of 88 lactating women in Singapore who received two doses of BNT162b2 vaccination (Pfizer/BioNTech), whereby outcomes of mother-child dyads within 28 days after the second vaccine dose were determined through a structured questionnaire. Minimal effects related to breastfeeding were reported in this cohort; 3 of 88 (3.4%) women had mastitis with 1 of 88 (1.1%) women experiencing breast engorgement. We report an incidence of lymphadenopathy in our cohort at 5 of 88 (5.7%). Reassuringly, there was no change in reported breastmilk supply after vaccination. The most common side effect was pain/redness/swelling at the injection site, which was experienced by 57 of 88 (64.8%) women. There were no serious adverse events of anaphylaxis and hospital admissions. No adverse symptoms were reported in 67 of 88 (76.1%) breastfed children.

**What’s known on this subject:** Two studies reported no serious adverse effects in both mother-child dyads after mRNA COVID-19 vaccination in mothers. Up to 61.9-67% lactating women experienced minor side effects.

**What this study adds:** We report an incidence of lymphadenopathy in our cohort at 5.7% as opposed to 0.3% from the Pfizer-BioNTech COVID-19 trial. Reassuringly, there was no change in reported milk supply after vaccination. Minimal effects related to breastfeeding were reported in this cohort; 3 (3.4%) women had mastitis with 1 person experiencing breast engorgement. The most common side effect was pain/redness/swelling at the injection site at 64.8%, which was experienced by 57 of 88 (65%) women. No adverse symptoms were reported in the breastfed children.

## INTRODUCTION

As clinical trials for the Pfizer/BioNTech mRNA COVID-19 vaccine, BNT162b2, did not include subjects who were breastfeeding, there is limited data on outcomes of breastfeeding mother-child dyads and effects on maternal milk production after vaccination.

Vaccination of lactating mothers is beneficial for both mother and infant, through protection of mothers from Covid-19, and transfer of passive immunity via milk to the infant.

In two previous studies,[1, 2] up to 61.9-67% lactating women vaccinated with either of the two mRNA COVID-19 vaccines (BNT162b2 by Pfizer-BioNTech and mRNA-1273 by Moderna) experienced minor side effects.

There have been anecdotal reports of mastitis and reduction in milk supply. However, there has been no systematic investigation into these effects. For clinicians to advise lactating women appropriately on COVID-19 vaccination, it is important to systematically quantify the incidence of local and systemic adverse events in this population.

As most of the lymphatic drainage of the breast is to the axillary lymph nodes, we hypothesize that BNT162b2, a COVID-19 mRNA vaccine, could result in an increased incidence of mastitis and/or breast engorgement with axillary lymphadenopathy after COVID-19 vaccination.

Hence, our aims were to determine: (1) solicited adverse effects such as axillary lymphadenopathy, mastitis and/or breast engorgement which are unique to lactating women, and (2) systemic and local adverse effects of COVID-19 mRNA vaccine on mothers and potential effects on their breastfed infants.

## MATERIALS AND METHODS

This is a prospective cohort study of lactating women in Singapore who received two doses of BNT162b2 vaccination (Pfizer/BioNTech). This study was approved by the National Healthcare Group Institutional Review Board (Gestational Immunity For Transfer GIFT-2: DSRB Reference Number: 2021/00095). The study protocol was registered at clinicaltrials.gov (<u>NCT04802278</u>).

Breastfeeding women who opted for BNT162b2 vaccination were invited to participate in the study via advertisements in healthcare facilities and social media. Informed consent was obtained from participants. All participants received 2 doses, 21 days apart, of 30ug (0.3ml volume) of BNT162b2 (Pfizer/BioNTech) vaccine intramuscularly in the deltoid region of their preferred arm. Demographics, past medical history, and clinical outcomes of mother-child dyads within 28 days after the second vaccine dose were determined through a structured questionnaire (Appendix A - Supplemental Material). In this study, only vaccine responses after second dose were studied.

## RESULTS

### Demographics of lactating women and their infants

Ninety-two pairs of women and infant dyads were recruited into the study. There were 4 dropouts. We report the results for the remaining 88 (92%) lactating women and their children who were enrolled in the study between February 2021 and March 2021. Table 1 shows demographics and clinical characteristics of the mother-child dyads. None of the women reported prior COVID-19 infection. All were non-smokers.

**Table 1:** Demographics and clinical characteristics of mother-child dyads

| Clinical characteristics | Lactating women who received 2<sup>nd</sup> dose of Pfizer-BioNTech COVID-19 vaccine (n=88) |
| --- | --- |
| Age, mean (SD), years | 33.2 (± 3.3) |
| Vaccinees' comorbidities, number (%)<br>- Asthma<br>- Allergic rhinitis<br>- Alpha thalassemia minor<br>- Thyroid cancer | 4 (5.3)<br>1 (1.3)<br>2 (2.7)<br>1 (1.3) |
| Ethnicity, number (%)<br>- Chinese<br>- Malay<br>- Indian | 75 (83.2)<br>6 (6.8)<br>2 (2.3) |
| - Others | 3 (3.4) |
| Type of feeds, number (%) |  |
| - Breastfeeding only | 71 (80.7) |
| - Breastfeeding and infant formula feeding | 17 (19.3) |
| Breastfed within 72 hours after COVID-19 vaccination, number (%) | 67 (76.1) |
| Age of child, mean (SD), months | 10.0 (± 5.2) |

### Reported adverse effects in lactating women after BNT162b2 vaccination (Pfizer/BioNtech)

#### Reported maternal lymphadenopathy, mastitis and human milk production

Five of 88 (5.7%) women experienced axillary or neck lymph node swelling. Three of 88 (3.4%) women reported mastitis. Another woman reported breast engorgement which resolved after 24 hours without a change in milk supply. No participant reported a change in milk supply (i.e., increased or decreased). One woman reported a transient bluish-green tinge to her milk colour after her first vaccine dose but not after her second dose.

#### Reported maternal-child local, systemic and/or serious adverse effects

Majority of the mothers reported local symptoms. The most common side effect was pain/redness/swelling at the injection site at 64.8%, which was experienced by 57 of 88 women. There were no serious adverse events of anaphylaxis and hospital admissions. 5.7% (5 of 88 women) experienced no symptoms. Detailed reporting can be found in Table 2.

**Table 2:** Reported symptoms of lactating women who received BNT162b2 vaccination

|  | **Lactating women who received**<br>**2<sup>nd</sup> dose of Pfizer-BioNTech**<br>**COVID-19 vaccine (n=88)** |
| --- | --- |
| Pain/redness/swelling at the injection site, number (%) | 57 (64.8) |
| Fever/chills, number (%) | 38 (43.2) |
| Nausea, number (%) | 1 (1.1) |
| Headache/ muscle ache/ joint pain, number (%) | 52 (59.1) |
| Fatigue, number (%) | 54 (61.4) |
| Lymphadenopathy (neck, axillary) number (%) | 5 (5.7) |
| Mastitis after vaccination, number (%) | 3 (3.4) |
| Breast engorgement, number (%) | 1 (1.1) |
| Anaphylaxis (%) | 0 |

No adverse events were reported for children who were breastfed after maternal vaccination, of which solicited responses were fever, rash, cough, behavioral change, vomiting and diarrhea.

## DISCUSSION

This is the first cohort study specifically addressing reported adverse events on breastfeeding and breastmilk production after maternal mRNA COVID-19 vaccination. We also reported other mother-child dyad outcomes to 28 days after completing second dose of vaccine.

We detected a incidence of lymphadenopathy in our cohort at 5.7% as opposed to 0.3% from the Pfizer-BioNTech COVID-19 trial.[3] Reassuringly, this was not associated with breast engorgement or mastitis. We postulate that lymphadenopathy in the draining lymph nodes from intramuscular injection site could be related to the greater immunogenicity associated with the COVID-19 mRNA vaccines. The reason for a higher incidence in our cohort could be a result of it being a solicited side effect as opposed to the original Pfizer-BioNTech COVID-19 trial whereby it was unsolicited.[3] Hence milder forms of lymphadenopathy might not have reported in the Pfizer-BioNTech COVID-19 trial.[3]

From a maternal standpoint, there was no appreciable increase in breast engorgement amongst vaccinated lactating women. There was also no reported maternal decrease in milk supply. 3.4% of the women in our cohort reported mastitis. This percentage is similar to the global epidemiological estimate of mastitis, which range from 2.5% to 20%.[4]

Interestingly, one woman reported a change in the color of milk expressed within 24 hours after her first dose of vaccination. The change in color of milk might not be related to the vaccine administration as it did not recur after the second dose of her vaccination. It has been reported that colour of human milk can vary widely and unusual milk color could be related to maternal dietary changes.[5]

Reassuringly, there were no short-term adverse effects reported in the children of 67 lactating women who breastfed within 72 hours after BNT162b2 vaccination.

This study has expected limitations. The reported outcomes are subjective reports from study participants; for example, there was no quantification of milk supply post-vaccination with a relatively short follow-up duration. As with other self-reported questionnaire, there is a possibility of recall bias resulting in over- or underestimating the events reported. Lastly, vaccine related effects were elicited after the second dose but not for dose one.

## CONCLUSION

Our findings suggest that the BNT162b2 vaccination is well tolerated in lactating women and not associated with short term adverse effects in their breastfed infants.

These results lend further evidence to current international recommendations that lactating individuals can safely continue breastfeeding their children after receiving COVID-19 mRNA vaccines.

## Data Availability

The data is confidential but can be shared in an anonymised manner upon request.

## Author Contributorship

Jia Ming Low - Substantial contributions to the conception or design of the work, acquisition and interpretation of data for the work, drafting the work

Le Ye Lee - Substantial contributions to the conception or design of the work, analysis and interpretation of data for the work, revising it critically for important intellectual content

Yvonne Peng Mei Ng - Substantial contributions to the design of the work, acquisition and interpretation of data for the work, revising it critically for important intellectual content

Youjia Zhong - Substantial contributions to the design of the work, acquisition of data for the work, acquisition and interpretation of data for the work, revising it critically for important intellectual content

Zubair Amin Substantial contributions to the conception or design of the work, analysis and interpretation of data for the work, revising it critically for important intellectual content

All authors gave final approval of the version to be published and are in agreement to be accountable for all aspects of the work in ensuring that questions related to the accuracy or integrity of any part of the work are appropriately investigated and resolved

## Conflict of interest disclosures

All authors declare no financial relationships with any organizations that might have an interest in the submitted work in the previous three years and no other relationships or activities that could appear to have influenced the submitted work. The preprint of the article is posted on https://www.medrxiv.org/

## Funding

J.M. Low received funding from KTP – NUCMI and National University Health System Pitch for Funds to conduct this research.

## Data availability

The data is confidential but can be shared in an anonymised manner upon request.

## Ethical approval and patient consent

This study was approved by the Institutional Review Board (Gestational Immunity For Transfer GIFT-2: DSRB Reference Number: 2021/00095). Informed consent was obtained from participants

## Acknowledgements

The authors would like to thank research coordinator Miss Regina Chua Xin Yi for helping with data collection and Dr Dimple Rajgor for helping with reviewing, formatting and in submission of the manuscript for publication. We would also wish to express our sincere gratitude to the mothers who donated their time and efforts to science.

## Supplemental Material: Structured questionnaire

## Appendix A

1. What is your date of birth?
2. Which ethnic group do you belong to?
  a. Chinese
  b. Malay
  c. Indian
  d. Caucasian
  e. Others
3. Are you a smoker? Yes/No
4. Any past medical history? Yes/No; if Yes, what is your past medical history
5. When was your first dose of COVID-19 vaccine?
6. When was your second dose of COVID-19 vaccine?
7. Did you have any vaccine-related side effects after the second dose of COVID-19? (Tick all that apply.)
  a. Pain, redness, swelling at the injection site
  b. Fever, chills
  c. Headache, muscle ache, joint pain
  d. Fatigue/tiredness
  e. Lymph node swelling (i.e. lumps) at neck or at armpits
  f. Allergic reactions/anaphylaxis
  g. No reaction to the vaccine
  h. Others
8. Did you get mastitis (i.e. painful breast swelling +/- fever) post vaccination? Yes/No
9. Did you have any breast-related complications/change in milk supply?
10. What is the date of birth of the child whom you are breastfeeding?
11. Please tick the following options as appropriate:
  a. Baby was fed breastmilk within 72 hours after COVID-19 vaccination and remained healthy
  b. Baby was fed breastmilk within 72 hours after COVID-19 vaccination and had an allergic reaction (e.g. rash/cough/difficulty breathing/behavioural change/fever/vomiting/diarrhoea)
  c. Baby was not fed breastmilk after COVID-19 vaccination
  d. Others
12. Please provide us with further feedback, if any

## Notes

### Competing Interest Statement

The authors have declared no competing interest.

### Clinical Trial

NCT04802278

### Author Declarations

Ethical approval was given and approved by the National Healthcare Group Institutional Review Board (Gestational Immunity For Transfer GIFT-2: DSRB Reference Number: 2021/00095).

